# Enveloped Virus Inactivation on Personal Protective Equipment by Exposure to Ozone

**DOI:** 10.1101/2020.05.23.20111435

**Authors:** Emmeline L. Blanchard, Justin D. Lawrence, Jeffery A. Noble, Minghao Xu, Taekyu Joo, Nga Lee Ng, Britney E. Schmidt, Philip J. Santangelo, M.G. Finn

**Author notes:** correspondence: Prof. M.G. Finn.

## Abstract

Ozone is a highly oxidizing gas easily generated from atmospheric oxygen with inexpensive equipment and is commonly used for the disinfection of municipal water, foods, and surfaces. We report tests of the ability of ozone to inactivate enveloped respiratory viruses (influenza A virus and respiratory syncytial virus), chosen as more easily handled surrogates for SARS-CoV-2, on N95 respirators and other personal protective equipment (PPE) commonly used in hospitals. At 20 ppm, an ozone concentration easily achieved by standard commercial equipment, the viruses were inactivated with high efficiency as long as the relative humidity was above a threshold value of approximately 50%. In the absence of humidity control, disinfection is more variable and requires considerably longer exposure under relatively dry conditions. This report extends the observations of a previous publication (http://doi.org/10.1080/01919510902747969) to hospital-relevant materials and provides additional details about the relationship of humidity to the antiviral activity of ozone. Home CPAP disinfection devices using ozone can provide effective results for individuals. Ozone did not appear to degrade any of the materials tested except for elastic bands if strained during treatment (such as by the pressure exerted by stapled attachment to N95 respirators). The filtration efficiency of N95 respirator material was not compromised. Overall, we recommend exposures of at least 40 minutes to 20 ppm ozone and >70% relative humidity at ambient temperatures (21-24°C) for 4-log (99.99%) reduction of viral infectivity on a variety of PPE, including gowns, face shields, and respirators. Shorter exposure times are likely to be effective under these conditions, but at the risk of some variability for different materials. Higher ozone concentrations and higher humidity levels promoted faster inactivation of viruses. Our work suggests that ozone exposure can be a widely accessible method for disinfecting PPE, permitting safer re-use for healthcare workers and patients alike in times of shortage.

## Introduction

Under conventional circumstances in high-resource environments, personal protective equipment (PPE), necessary to protect wearers from a variety of harmful substances such as biological waste, bacteria, and viruses,^1^ is often designed to be discarded after a single use.^2^ However, high demand during the SARS-CoV-2 pandemic has led to shortages in PPE and created the need for its safe reuse.

While a great deal has been published on the disinfection of materials and surfaces for a wide variety of industries, only a few reports exist that are focused on the effectiveness of disinfectants on PPE.^3-5^ Numerous possibilities exist, including alcohols, heat, and ultraviolet light.^5-9^ We focus here on ozone, an easily generated and highly oxidative gas that is widely employed in a variety of applications, including for industrial-scale food disinfection^10^ and municipal wastewater treatment.^11,12^ To our knowledge, only one report has recently appeared concerning the use of ozone for the disinfection of PPE, focusing on N95 respirators.^13^ We describe a similar study here, extended to other relevant PPE materials and exploring the role of humidity in the effectiveness of ozone treatment.

SARS-CoV-2 is an enveloped virus, as are all other coronaviruses and many other respiratory viruses.^14^ We were surprised to find that the vast majority of the disinfectant products recommended by the U.S. Environmental Protection Agency for use against coronavirus contamination have been tested only on nonenveloped viruses (those lacking an outer lipid membrane).^15^ Similarly, ozone disinfection has also been tested more frequently on non-enveloped viruses than enveloped ones,^9,16-21^ and only rarely has the deactivation of coronaviruses by multiple methods been studied.^22,23^ We wished to explore the parameters for ozone deactivation of a respiratory virus that would serve as a reasonable surrogate for the highly infectious SARS-CoV-2. We chose influenza A virus (IAV; strain A/WSN/33) as our primary test pathogen, and human respiratory syncytial virus (RSV; strain A2) as a secondary target, both of which are handled under BSL-2 (biosafety level 2) conditions. As summarized in **Table 1**, these pathogens are similar in form and function to SARS-CoV-2, although not without some differences.

**Table 1.** Pathogens used as surrogates for SARS-CoV-2

|  | **Influenza virus A**<br>**A/WSN/33** | **RSV A2** | **SARS-CoV-2** |
| --- | --- | --- | --- |
| **Particle size** | spherical ≈100 nm<br>diameter, filamentous<br>>300 nm | spherical pleomorphic<br>≈200 nm, filamentous<br>= several microns | elliptical, pleomorphic<br>≈100 nm diameter |
| **Genome** | (-)ssRNA, 13.6 kB total,<br>8 segments (0.9-2.3 kB) | (-)ssRNA, 15 kB | (+)ssRNA, ≈30 kB |
| **Human host**<br>**cells** | respiratory epithelium | respiratory epithelium,<br>ciliated cells | lung alveolar type 2<br>cells, others |

We explored three questions regarding ozone disinfection. (1) Does ozone successfully neutralize virions, such that they are no longer infectious? (2) Does the PPE mechanically survive ozone treatment? (3) Does ozone treatment negatively impact the proper protective function of the PPE, especially the filtration properties of N95 respirators? We examined these questions using several commercially available ozone treatment units marketed for use in other contexts. We hope this will be of value to potential users considering the feasibility of the safe reuse of PPE. A 2009 report described similar findings for the inactivation of 12 viruses, including seven enveloped viruses (influenza, herpes simplex virus, sindbis virus, yellow fever virus, vesicular stomatitis virus, vaccinia virus, and mouse coronavirus) when deposited on a hard surface.^24^

## Results and Discussion

### Assay Validation

In the literature, virus survival post-disinfection is typically measured by determining viral titer by plaque assays or TCID_50_ (median tissue culture infectious dose) on permissive cells, or by measuring viral RNA loads via RT-qPCR.^18,25^ We sought to maintain viral infectivity as the primary readout, but to do so in a simpler and faster way. Accordingly, we employed the reporter strains WSN/33-PA-2A-NLuc and RSV-luc5, replication-competent influenza A virus and respiratory syncytial virus, respectively, modified to have a cleavable NanoLuc (NLuc) reporter on the PA protein (IAV) and a luciferase between the P and M genes (RSV).^26,27^ Overnight passage of any remaining virus after ozone treatment on MDCK cells (for IAV) or HEp-2 cells (for RSV) followed by cell lysis and NLuc or luciferase signal quantification provided a fast and convenient readout of viral protein load, and thereby of viral replication.

Testing with known concentrations of IAV and RSV demonstrated the dynamic range of the NLuc reporter assay (factors of 10^6^ or greater in virus dilution) to exceed those of RT-qPCR and plaque assays (10^4^-fold range) in our hands (**Figure 1a-c, f-g**). Asterisks in **Fig. 1b** indicate dilutions where large cytopathic effects were observed, such that plaques were uncountable. We also treated representative IAV samples with ozone in a standard manner (see below), and analyzed half of the samples by RT-qPCR and the other half with the NLuc reporter assay. As shown in **Fig. 1d-e**, the observed trends were similar; these results with a small number of samples in each group also illustrate the general observation that the NLuc assay provided somewhat smaller variance among replicates.

**Figure 1.**
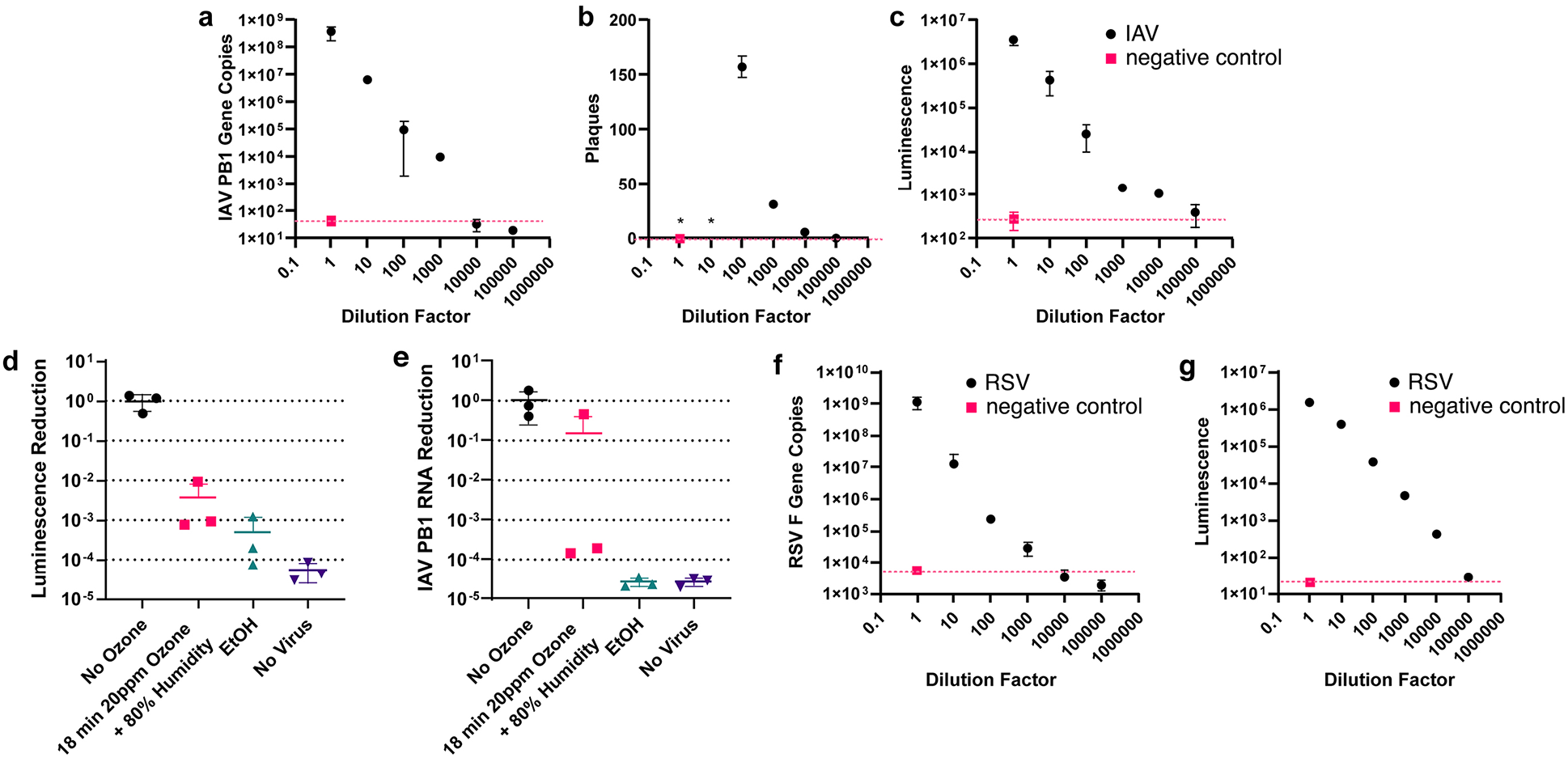
Comparison of assay methods for virus viability. Variation in IAV concentration assayed by (a) RT-qPCR, (b) plaque assay, (c) NanoLuc. (d,e) Treatment of IAV on N95 material with ozone, assayed by (d) NanoLuc and (e) RT-qPCR. Variation in RSV concentration assayed by (f) RT-qPCR, (g) Luc.

### Effects of exposure time, humidity, and ozone concentration on virus inactivation

We selected three types of materials in common use in healthcare environments for testing of virus deactivation by ozone: cloth face masks, Tyvek (spun high-density polyethylene) fabric used in disposable gowns and PAPR (powered air purifying respirator) hoods, and N95 respirators (Supplemental Information, **Table S1**). In each case, a measured amount of virus was applied to small (1 cm square) pieces of material, allowed to dry, and then subjected to disinfection. Any remaining virus was recovered by immersion of the treated material in growth media, followed by passage through susceptible cells in culture. For multi-layered materials such as the N95 respirators, the virus solution did not remain on the surface layer, but rather were absorbed into the material. For negative disinfection controls, the same types of samples were treated with virus, left to dry, and were kept at room temperature without treatment. Virus-laden samples sprayed with 70% ethanol were allowed to dry and served as positive disinfection controls. Minimum assay response was defined by the signal generated by processing of materials not treated with any virus. In each experiment, 4-6 replicates of each sample type were used. To account for experiment-to-experiment variation in absolute values, results are reported as % reduction vs. ethanol treatment, which consistently provided more than 10^4^-fold reduction in viral infectivity.

Ozone exposure was performed using three commercial instruments. Two (manufactured and provided by Global Ozone Innovations and Zono Technologies) were cabinets used primarily to disinfect sports and playground equipment in schools, daycare centers, and the like. The third was a disinfection device for home PAP (positive airway pressure) equipment (VirtuCLEAN, provided by Healthcare Logiix Systems). These devices were obtained by virtue of personal contacts with company representatives, not because of any particular technical features, and we believe they are representative of the types of equipment that can be widely obtained. Ozone and humidity were monitored with built-in or externally-introduced sensors. Hardware details are given in **Tables S2 and S3**; **Figure S1** shows representative time courses of changes in ozone and humidity levels through a typical run. One of the instruments (made by Zono Technologies) was able to simultaneously control ozone concentration, humidity, and temperature, and so this was used for most of the experiments investigating changes in these parameters.

A standard ozone concentration of approximately 20 ppm was produced by all of the units. Under humid conditions (80% relative humidity, RH, at room temperature), effective inactivation of influenza virus (>70% of the ethanol control) was achieved even at the shortest exposure time tested (18 min), with a tendency to increase at longer times (up to 90 min), as shown in **Figure 2a**. In contrast, when the humidity was not controlled, disinfection was much more variable and required up to 4 h of ozone treatment to achieve results comparable to 70% ethanol for all three materials tested (**Fig. 2b**).

**Figure 2.**
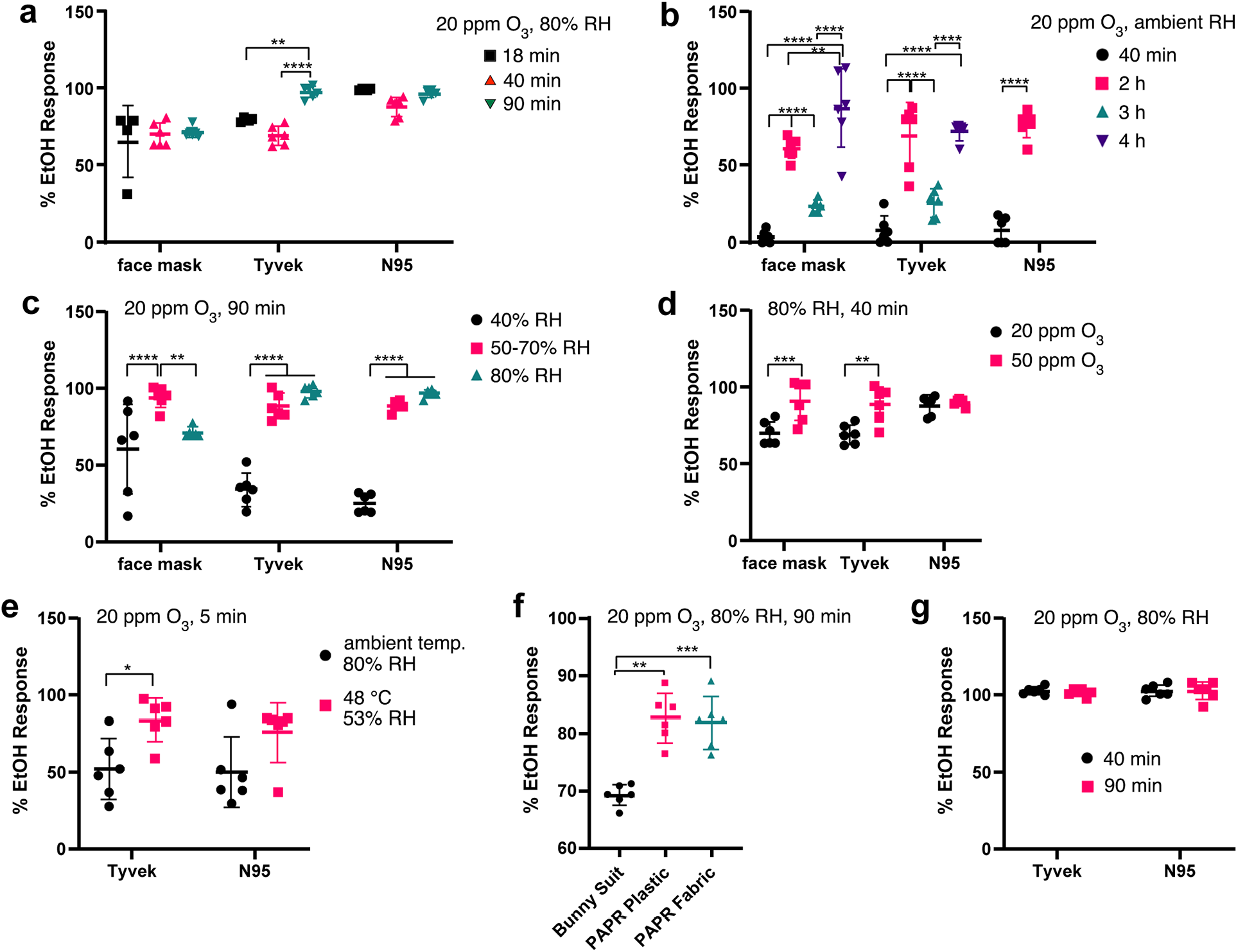
(a-f) Inactivation of influenza A virus by ozone under various conditions. Vertical axis = decrease in IAV infectivity relative to the results obtained from saturation of virus-deposited materials with 70% ethyl alcohol relative to samples receiving no disinfection treatment. (e) 80% RH at 24 °C = approx. 17.4 g/m^3^ water vapor; 53% RH at 48°C = approx. 40 g/m^3^. (g) Inactivation of RSV by ozone. RH = relative humidity. All experiments were performed at room temperature (24 ± 1 °C) unless otherwise indicated. 2way ANOVAs with Tukey’s multiple comparison tests were performed for the data in panels a, c-e, and g. A 2way ANOVA with a Tukey’s multiple comparison test was run for panel b, except for N95 samples, for which a t-test was performed. A Brown-Forsythe and Welch ANOVA with a Dunnet’s T3 multiple comparisons test was run for panel f. * p < 0.02, ** p < 0.01, *** p < 0.001, and **** p < 0.0001.

The results of a series of experiments controlling relative humidity (**Fig. 2c**) demonstrated the critical nature of this parameter. A clear threshold was observed for all three materials, with 40% RH being ineffective and 50-70% RH giving maximum performance. These results are quite similar to those reported recently by Duchaine and colleagues on the deactivation of aerosolized nonenveloped viruses (phage and norovirus examples).^21^ Increasing to 80% RH provided no significant additional benefit. Increasing ozone concentration from 20 ppm to 50 ppm gave a predictably better response, but the lower concentration was nevertheless quite effective (**Fig. 2d**). Higher temperature (48°C vs. room temperature) under humid conditions allowed for significant reduction of IAV infectivity on both Tyvek and N95 material in only 5 minutes, but this of course is less convenient to perform in practice (**Fig. 2e**). In this experiment, a higher water vapor concentration was maintained at the higher temperature even though the relative humidity value was lower. In contrast, increasing the temperature to 45°C while maintaining humidity at a relatively low level did not significantly enhance the rate of virus inactivation by ozone (**Figure S2**).

Extended (90 min) ozone treatment under humid conditions also successfully inactivated IAV on samples of Tyvek from a standard disposable coverall (“bunny suit”) and a hospital PAPR hood, as well as the polycarbonate PAPR face shield (Fi**g. 2f**). Lastly, RSV was completely deactivated by ozone under the same conditions, after either 40 or 90 minutes exposure (**Fig. 2g**). Therefore, if IAV and RSV are representative, these results suggest that ozone treatment at 20 ppm or greater, 70% or greater relative humidity at room temperature, and for at least 40 minutes should reliably inactivate enveloped viruses on a variety of materials of use in medical PPE.

### Effects of ozone on materials properties

We exposed larger swaths of Tyvek fabric from various sources, as well as cloth masks, N95 respirator material, and polycarbonate face shields to multiple cycles of ozone exposure (20 ppm) adding up to two hours or more. With one important exception, no perceptible changes occurred in any material, including goggle transparency, the integrity of seams and glued components, and the appearance and function valve assemblies and other plastic components (such as zippers). While we did not perform quantitative fit testing of the respirators post-treatment, no deformations were observed following any of the ozone treatment variations. A summary of mechanical assessments is provided in **Table S4**. Quantitative rheological measurements were not performed.

Only the elastic headbands of some N95 respirators were found to be degraded by ozone treatment, and only when the material was strained during exposure, either by function of a stapled attachment to the respirator or by tying off in a stretched state in the ozone cabinet (**Figure S3**). The degradation of strained rubber upon exposure to ozone has been previously described,^28^ presumably due to scission of C=C double bonds in the polymers which results in cracking orthogonal to the strain. We found that even a short exposure of strained elastics to 20 ppm O_3_ (15 minutes, less than generally required for viral inactivation) often resulted in compromised headband integrity or complete failure.

A subset of NIOSH-approved N95 respirators cleared by the Food and Drug Administration (FDA) are referred to as Surgical N95 Respirators.^29^ While we were unable to obtain samples of these FDA-approved respirators, many are manufactured with stapled headband attachments. This presents a significant practical barrier to the use of ozone to disinfect N95 respirators. However, headbands made of thermoplastic elastomer or polyisoprene with polypropylene overbraid can be attached by ultrasonic welding (such as the 3M 8210 and ZKG9501 KN95 respirators that we were able to obtain for this study). These headbands showed no degradation upon several hours of repeated ozone treatments (**Figure S4**), and so we surmise that welded attachments do not induce significant elastomer strain at the point of attachment.

### N95 particle filtration efficiency

Full-thickness sections of N95 respirator material before and after extensive ozone exposure were tested for filtration ability using sodium chloride aerosols in a manner closely analogous to the NIOSH N95 respirator penetration testing guidelines.^30^ No substantial loss of filtration performance was found for these ozone-treated materials (**Figure S5**, **Table S5**).

## Additional experiments and conclusions

Ozone is readily and inexpensively generated and has a long history of disinfection use against bacteria, fungi, and other viruses. As a highly oxidizing and diffusible species, it causes damage to lipid membranes,^31^ amino acids,^32,33^ and polynucleotides;^20,34-40^ an early report on a nonenveloped virus (poliovirus) identified genomic damage as the most likely mechanism of viral inactivation.^41^

We established here that ozone is highly effective in the inactivation of enveloped respiratory viruses that serve as surrogates for coronaviruses, as long as the relative humidity during ozone exposure is approximately 50% or greater at room temperature. Although not presented above because of inadequate numbers of replicates or controls, we made several additional observations that may be useful to others. (i) We found that the deactivation of viruses could be enhanced by the simple expedient of including water-saturated paper towels in the sample space of the personal PAP cleaning device and the larger Global Ozone Innovations (GOI) cabinet. (ii) A brief test of plastic tubing used in a standard hospital ventilator showed highly effective virus deactivation under the same conditions found effective for Tyvek, but care must be taken to make sure that ozone can reach throughout the interior of the ventilator tube. Just placing a coiled length of ventilator tubing inside an ozone cabinet may not ensure adequate air flow for this purpose. (iii) We also had the opportunity to test the performance of a trailer equipped by GOI to disinfect a large number of garments or other pieces of equipment in a single run. This unit used 16 corona discharge generators to disinfect a volume of 30 cubic meters, probably reaching ozone concentrations of hundreds of ppm. Our results were highly variable due to variations in ambient relative humidity, following closely on the trends described above for more highly controlled devices.

In addition to the aforementioned studies of nonenveloped^21^ and enveloped^24^ viruses, the key role of relative humidity in ozone reactivity has additional precedent in the literature. Humidity has been found to be an important accelerant of ozone reactivity with small organic molecules in aerosol particles, for reasons having to do with substrate solubility at the particle-gas interface^42^ and other surface properties and chemistries.^43,44^ Similarly, lignin degradation by ozone is sensitive to moisture for complex reasons,^45^ On the other hand, relative humidity was not found to be a strong contributing factor to the reactivity of ozone with other kinds of surfaces, such as inorganic oxides.^46,47^ We suggest that humidity may play at least two potential roles in rendering ozone more damaging to enveloped viruses: (a) water may promote the generation of highly reactive hydroxyl radicals from ozone,^48,49^ and (b) more humidity may plasticize surfaces or otherwise make it easier for ozone to interact with surface-bound species, such as by diffusion to reach the viruses.

As noted above, the major impediment to the use of ozone for the decontamination of large numbers of N95 respirators is damage caused to elastic headbands when secured by staples. This would seem to require detachment and reattachment of the elastic before and after ozone treatment (the elastic can be treated with ozone as long as it isn’t stretched), or by re-engineering of the attachment method such as by replacing the bands with cloth ties. However, scalable humid ozone treatment does seem to be a robust potential solution for many other materials, in addition to airborne viruses.^21^

## Methods

### Description of fabric sample swatches

Fabric samples for viral inoculation were prepared by cutting 1 cm × 1 cm sample swatches from four different materials: surgical face masks, Tyvek (disposable gown), N95 respirators, bunny suits, and PAPR hoods (both the fabric and plastic face shield).

### Cell lines and Viruses

MDCK (ATCC) and HEp-2 (ATCC) cells were maintained in DMEM + 10% fetal bovine serum and 1% pen-strep at 5% CO2 and 37°C. H1N1 Influenza A WSN/33-PA-2A-NLuc (BEI) and RSV-luc5 (Viratree) reporter strains were obtained from the indicated suppliers, and designated here as “IAV” and “RSV,” respectively.

Viral stocks were prepared as follows. IAV was propagated in MDCK cells, where virus was added to cells at a multiplicity of infection (MOI) of 0.01 for 45 minutes before being removed and replaced with virus growth medium (DMEM with 1% pen-strep, 2 mM L-glutamine, 0.2% bovine serum albumin, and 25 mM HEPES buffer). Virus was harvested when 90% of the cytopathic effects were visualized. Media was collected, spun down at 300×g for 15 minutes to remove cell debris and supernatant was aliquoted. RSV was propagated in HEp-2 cells, where virus was added to cells at a MOI of 0.1 for 1 h before adding virus growth medium (DMEM with 10% fetal bovine serum and 1% pen-strep) to the inoculum. Cell-associated virus was harvested by scraping the cells when 90% of cytopathic effects were visualized. Virus-containing media was then vortexed and aliquoted for use.

### Sample preparation and disinfection

Virus solutions (50 μL) in growth media at ~10^5^ PFU/ml were deposited by pipette onto 1 cm × 1 cm pieces of each candidate material, as evenly as possible over at least half of the surface area. The samples were allowed to dry in air at room temperature for 30 minutes and were transported to the disinfection unit in plastic cases. For initial treatment experiments, samples were left inside open cases during ozone exposure. To better approximate the treatment of actual intact PPE garments and materials, most subsequent samples were pinned to cotton shirts and hung in the ozone cabinets so as to allow air/ozone access to both sides of the sample swatch. For the small zippered pouch of the VirtuCLEAN PAP cleaning device, the samples were pinned to an N95 respirator placed inside the pouch.

No-disinfection control samples remained at room temperature dry and untreated, while ethanol controls were sprayed with 70% ethanol and then allowed to dry and remain at room temperature for the duration of the disinfection procedure. After treatment (or control non-treatment), fabric samples were immersed in 300 μL of virus growth media (for IAV, DMEM with 1% pen-strep, 2 mM L-glutamine, 0.2% bovine serum albumin, and 25 mM HEPES buffer; for RSV, DMEM with 10% fetal bovine serum and 1% pen-strep) for 30 minutes at room temperature in the top of a 0.65 μm filter. Each sample was then spun at 12000×g for 2 minutes to extract virus/media, designated as “virus inoculum.” For virus applied to hard plastic surfaces, the surface was swabbed with a sterile cotton swab dipped in virus growth media. The cotton swab was then incubated in 300 μL of virus growth media for 30 minutes at room temperature, followed by removal of the swab and centrifugation at 12000×g for 2 minutes. MDCK cells (IAV) or HEp-2 cells (RSV) plated in a 24 well plate at confluency were washed once with 1×PBS and then incubated with 100 μL of virus inoculum for 1 hour at 37°C, being rocked every 15 minutes. After 1 hour, the inoculum was removed and the cells were washed once with 1×PBS. 500 μL of virus growth media was added to cells, which were then incubated at 37°C overnight before analysis.

### Plaque Assay

MDCK cells were plated in 6-well plates until confluent. Cells were washed twice and incubated with 200 μL of virus inoculum, diluted in DMEM + 0.1% trypsin-TPCK, for 1 h at 37°C, being rocked every 15 minutes. The inoculum solution was then removed, the cells were washed once, and then were incubated at 37°C with an Avicel-DMEM overlay media containing a 1:1 ratio of 2.4% Avicel (FMC Biopolymer RC-481) and DMEM, supplemented with 0.2% BSA, 1% pen-strep, 2 mM L-glutamine, 0.1% trypsin-TPCK, and 25 mM HEPES buffer. After 3 days, the overlay was removed and cells were fixed in 4% paraformaldehyde for 30 minutes, before being stained with crystal violet (0.1% in 20% ethanol) for 30 minutes. After the crystal violet was removed, the monolayer was air dried and plaques were counted.

### RT-qPCR

The supernatant was removed and the cells were washed and then processed with an RNAeasy PLUS RNA extraction kit (Qiagen) to extract RNA. cDNA was then generated from isolated RNA with a High-Capacity cDNA Reverse Transcription Kit (ThermoFisher). RT-qPCR was then performed for IAV PB1, with Taqman probe (ACACGAGTGGACAAGCTGACACAA) and primers (ATCTTTGAGACCTCGTGTCTTG and CAGCAGGCTGGTTCCTATTTA) or for RSV F, with Taqman probe (TGCCATAGCATGACACAATGGCTCCT) and primers (AACAGATGTAAGCAGCTCCGTTATC and CGATTTTTATTGGATGCTGTACATTT) (ThermoFisher).

### NLuc Reporter Assay

The cell media was removed and the cells were washed once with 1×PBS. 250 uL of Nano-Glo Luciferase Assay Reagent (Promega) (1:100 ratio of Nano-Glo Luciferase Assay Substrate:Nano-Glo Luciferase Assay Buffer) was added per well and incubated for 3 minutes at room temperature. 100 uL of Nano-Glo Luciferase Assay Reagent was pipetted from each well as technical duplicates into a white 96 well plate and luminescence was quantified.

### Luciferase Reporter Assay

Supernatants were removed and cells were washed once with 1×PBS. 250 ul of Bright-Glo Luciferase Assay Reagent (Promega) was added per well and incubated for 3 minutes at room temperature. 100 ul of Bright-Glo Luciferase Assay Reagent was pipetted from each well as technical duplicates into a white 96 well plate and luminescence was quantified.

### Mechanical test methods

We treated various pieces of equipment for multiple cycles to assess mechanical tolerance of PPE to ozone treatment compared to control items. Items tested are listed in **Table S4**.

### Particle filtration efficiency testing methods

Seven different respirators were assessed at 20 ppm ozone, with minimum treatment durations ranging from 160 to 320 min. Sample material from each of the respirators that underwent mechanical tests were used to assess post-treatment particle filtration efficiency. Circular samples of 2.5 cm diameter were punched from each respirator and placed into a filter holder. Aerosol size distribution and number concentration with and without the sample were measured by a Scanning Mobility Particle Sizer (SMPS) system^50^ to determine particle filtration efficiency. The SMPS system consists of a Differential Mobility Analyzer (TSI 3080) and a Condensation Particle Counter (TSI 3775). Polydispersed aerosols were generated via atomization of 0.05 M sodium chloride solution. Experiments were conducted at 1.92 LPM. Two sets of testing were performed per each sample and averaged. We note that these tests were not completely equivalent to the NIOSH standard, and differences include but not limited to the following:

- Tested respirators were not pre-conditioned at 38°C and 85% relative humidity
- Aerosol mass concentration was substantially lower in this test
- Aerosol size distribution was skewed toward smaller size compared to NIOSH standard.

### Ozone treatment methods

From April 2 to April 24, 2020, we assessed four different ozone treatment devices from three manufactures, Global Ozone Innovations (GOI), Zono Technologies (ZT), and VirtuCLEAN (VC) (**Table S2**). These devices are designed to disinfect recreational or medical equipment, and have been largely evaluated against bacterial pathogens,^51^ but few studies have assessed the efficacy of these technologies against encapsulated viruses. The GOI and VC units rely on ozone alone, whereas the ZT cabinet also treats with elevated (80%) humidity via a water reservoir and ultrasonic humidifier. The GOI and VC devices generate ozone via corona discharge and the ZT cabinet, which additionally uses elevated humidity, uses 184 nm photolytic UV ozone generators because humidity reduces corona plate generation efficiency. When not otherwise noted, all experimental runs were performed at ambient temperature (ca. 24-25°C) and pressure.

All the devices tested use atmospheric oxygen to generate a nominal ozone concentration of approximately 20 ppm. Whereas the GOI and ZT cabinets control this by virtue of internal sensing and real-time regulation, the less expensive VC cleaner turns ozone generation on and off to give rise to cycles of ozone concentrations ranging from 10-30 ppm, averaging 17-20 ppm (**Figure S1**). To extend runs beyond the time limits imposed by the manufacturer’s hard-coded time limits, the VC device was simply restarted immediately at the end of each treatment cycle. Both GOI and ZT provided extensive remote or on-site engineering support to permit modification of time and concentration set points in support of these experiments.

The Zono Technologies cabinet and VirtuCLEAN device do not offer data logging and export, so a SPEC Sensors Digital O_3_ Sensor was placed inside the devices and used to log ozone concentration, temperature, and relative humidity levels during runs (see **Figure S1**). While these sensors are rated from 0-5 ppm O_3_, we observed they matched the readings from an EcoSensor SM-7 0-50 ppm range sensor in the ZT cabinet up to at least 45 ppm. These sensors, as well as the EcoSensor SM-7, required a warm up period for reliable operation.

Each instrument device uses a ventilation or ozone destruction cycle (reversing airflow through a catalyst) following treatment to return ozone concentrations back to safe levels. The ZT instrument passes the internal atmosphere continuously over a catalyst until ozone levels are reduced, and the VC unit completes a 5 minute vent through a catalyst (which worked well in our hands). All devices tolerated back-to-back runs over multiple hours with no change in ozone generation, and require no consumables chemicals or reagents, other than water.

## Data Availability

Raw data are given in the Supplemental Excel files.

## Acknowledgements

We are extraordinarily grateful to Mark Eades and Rik Kain (Global Ozone Innovations) and Walter Mann (Zono Technologies) for generously making their instruments and expertise available to us in timely fashion, and to Everette Webb and Kyle Miko of Healthcare Logiix Systems for two PAP cleaning units. This work was supported by the Georgia Institute of Technology and by grant funding from (i) the Center for Pediatric Nanomedicine at the Georgia Institute of Technology, Emory University, and Children’s Healthcare of Atlanta, (ii) NIH R01GM114561 (to P.J.S.) and (iii) NASA FINESST Fellowship 80NSSC19K1544 (to J.D.L.) and NASA grant 80NSSC18K1301 (to B.E.S.). J.A.N. acknowledges support in part by the NIH (5T32EB021962-02) and the NSF Graduate Research Fellowship program (DGE-1650044). T.J. and N.L.N. acknowledge support from the Center for the Science and Technology of Advanced Materials and Interfaces (STAMI) at Georgia Tech.

## Disclaimer

No financial support was received from any commercial entity and the findings reported here have not been reviewed or endorsed by any company. While our observations do not suggest the presence of hidden effects, there may be negative consequences to material integrity or performance resulting from ozone treatment that we have not identified.

## Supplemental Information

**Table S1.**
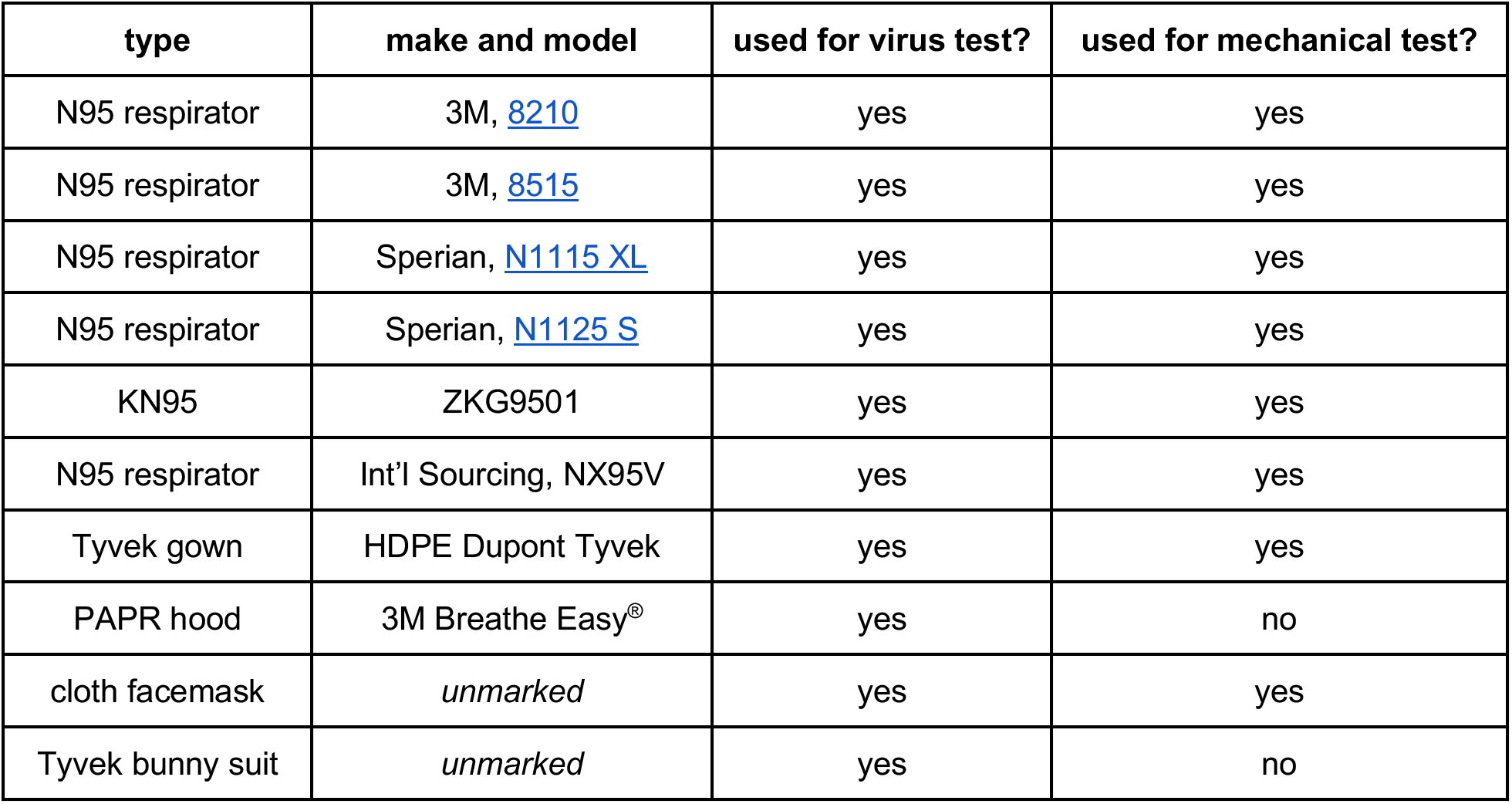
Materials used in assessments of virus inactivation, mechanical robustness, and filtration efficiency.

**Table S2.** Ozone treatment devices assessed.

| Device | Volume (m<sup>3</sup>) | Standard Time (m) | Standard [O<sub>3</sub>] (ppm) | Typical Humidity | O<sub>3</sub> generator | Control Sensor, Logging Sensor |
| --- | --- | --- | --- | --- | --- | --- |
| Global Ozone Decon-Zone 4201A Cabinet | 0.53 | 16 | 20 | ambient* | corona discharge | EcoSensor SM-7 0-20 ppm, recorded to SD card at 0.1 Hz |
| Global Ozone OT-100 Trailer | ~30 | < 99 | ≥ 20 | ambient* | corona discharge | EcoSensor SM-6 0-20 ppm, recorded via USB O<sub>3</sub> Sensor at 1 Hz** |
| Zono SC 1 Cabinet | 0.73 | 18 | 20 | 80% | deep UV | EcoSensor SM-7 0-50 ppm, recorded via USB O<sub>3</sub> Sensor at 1 Hz** |
| VirtuCLEAN 2.0 Waterless CPAP Cleaning Pouch | < 0.01 | 30 | 15-16 | ambient* | unknown | No concentration readout. Recorded via USB O<sub>3</sub> Sensor at 1 Hz** |

\*not controlled
\*\*SPEC Sensors Digital O<sub>3</sub> Sensor \(DGS-O3 968-042\)

**Table S3.**
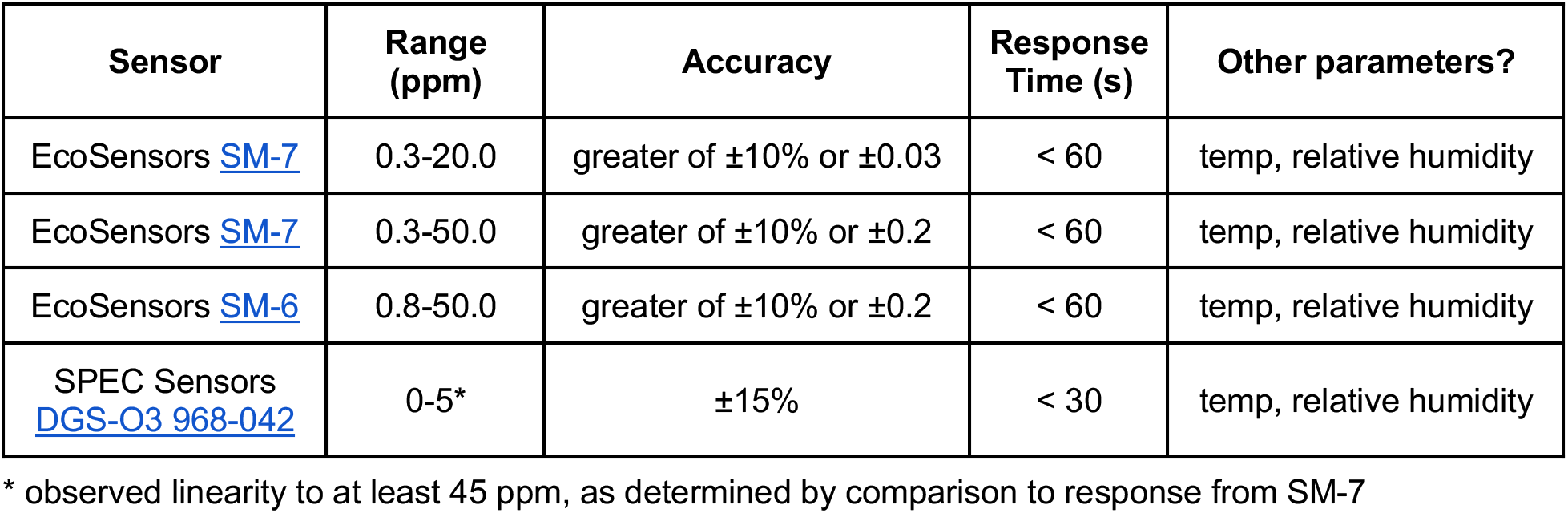
Characteristics of ozone, temperature, and humidity sensors used.

| Sensor | Range (ppm) | Accuracy | Response Time (s) | Other parameters? |
| --- | --- | --- | --- | --- |
| EcoSensors SM-7 | 0.3-20.0 | greater of ±10% or ±0.03 | < 60 | temp, relative humidity |
| EcoSensors SM-7 | 0.3-50.0 | greater of ±10% or ±0.2 | < 60 | temp, relative humidity |
| EcoSensors SM-6 | 0.8-50.0 | greater of ±10% or ±0.2 | < 60 | temp, relative humidity |
| SPEC Sensors DGS-O3 968-042 | 0-5* | ±15% | < 30 | temp, relative humidity |

\* observed linearity to at least 45 ppm, as determined by comparison to response from SM-7

**Figure S1.**
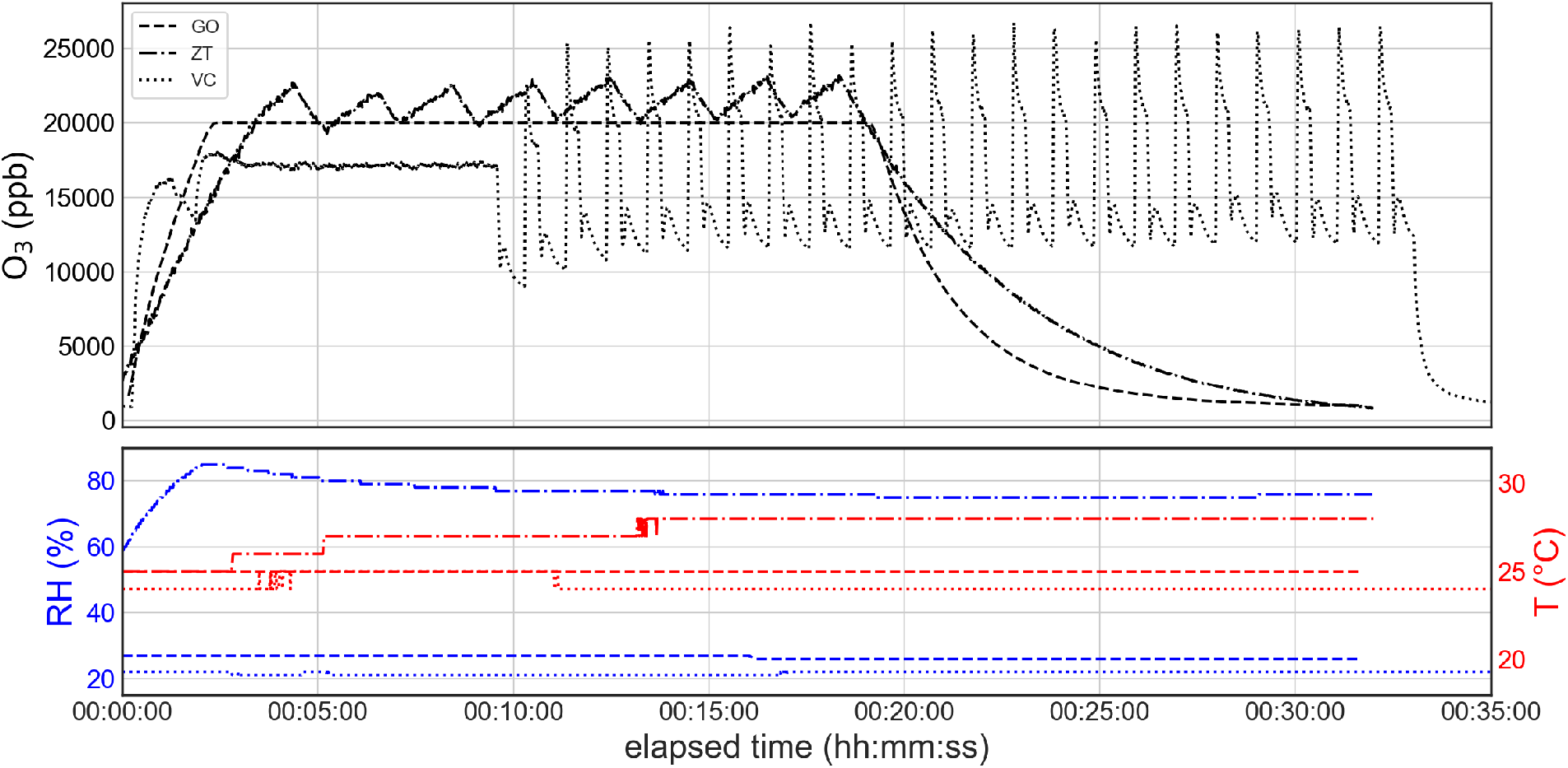
Comparison of ozone concentration, temperature, and humidity during a standard run cycle for each device. Line style convention for bottom plot follows from first plot. Also demonstrates these treatment devices do not exceed commonly recommended N95 respirator storage temperatures of < 30°C. GO = Global Ozone cabinet, ZT = Zono Technologies cabinet, VC = VirtuCLEAN portable PAP disinfection zippered pouch.

**Figure S2.**
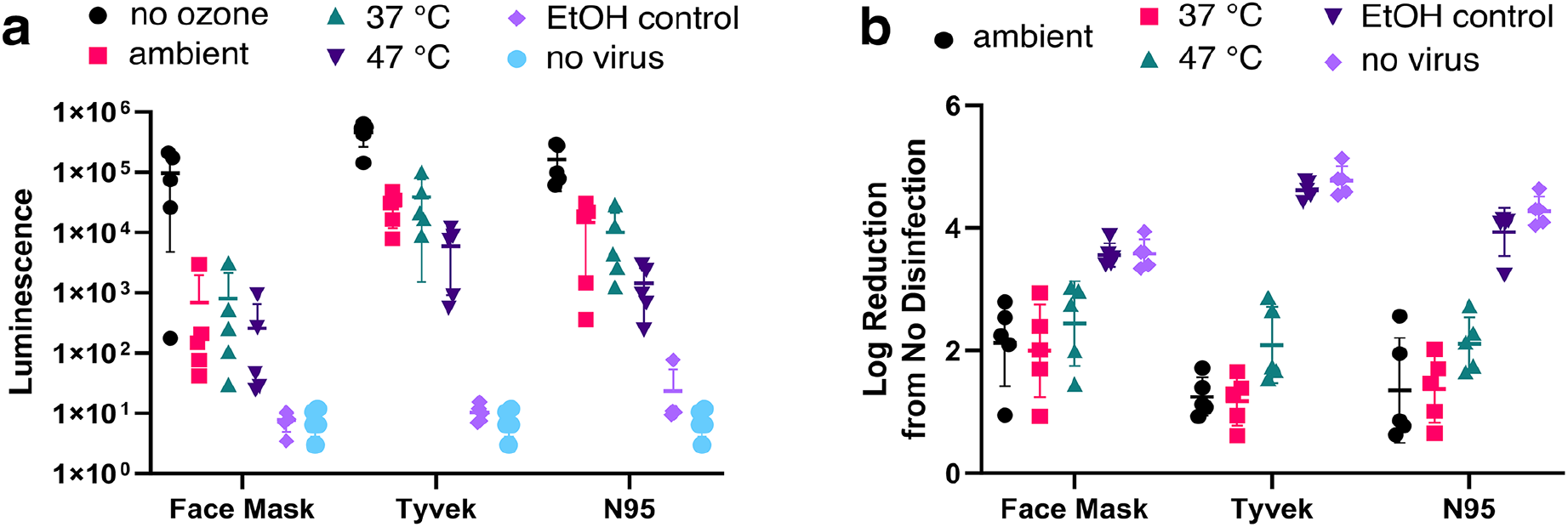
Inactivation of influenza A virus by ozone as a function of temperature, holding atmospheric moisture roughly constant. Here the data are displayed for illustrative purposes in terms of (a) observed luminescence from the NanoLuc assay and (b) reduction of viral infectivity derived from the data in panel (a). Approximate water vapor concentrations: ambient (25 °C), 14.3 g/m^3^ = 62% RH; 37 °C, 14.5 g/m^3^ = 33% RH; 45 °C, 17.7 g/m^3^ = 27% RH.

**Table S4.** Results of mechanical assessments.

| item | make and model | exposure time (20 ppm) | observations of appearance and mechanical properties |
| --- | --- | --- | --- |
| N95 respirator | 3M, 8210 | 320 m | no significant changes |
| N95 respirator | 3M, 8515 | 160 m | No changes in respirator material, elastic bands failed at staple attachment |
| N95 respirator | Sperian, N1115 XL | 320 m | No changes in respirator material, elastic bands failed at staple attachment |
| N95 respirator | Sperian, N1125 S | 230 m | No changes in respirator material, elastic bands failed at staple attachment |
| N95 respirator | KN95, ZKG9501 | 160 m | no significant changes |
| N95 respirator | NX95V | 160 m | No changes in respirator material, elastic bands failed at staple attachment |
| Surgical mask | *unmarked* | 320 m | no significant changes |
| clear polycarbonate (face shield, goggles) | *unmarked* | 160 m | no significant changes |
| Tyvek disposable gown | *unmarked* | 160 m | no significant changes |
| Tyvek PAPR hood fabric | 3M Breathe Easy® | 160 m | no significant changes |

**Table S5.** Results of particle filtration efficiency assessments.

| Manufacturer | Model | Duration of ozone exposure | Control <sup>a</sup> | Result <sup>b</sup> |
| --- | --- | --- | --- | --- |
| 3M | 8210 | 320 min | yes | pass |
| 3M | 8515 | 160 min | no | pass |
| Sperian | N1115 XL | 320 min | yes | pass |
| Sperian | N1125 S | 230 min | no | pass |
| KN95 | ZKG9501 | 160 min | no | unknown <sup>c</sup> |
| International Sourcing | NX95V | 160 min | no | unknown <sup>d</sup> |
| *unmarked* | surgical mask | 320 min | yes | pass <sup>e</sup> |

**Figure S3.**
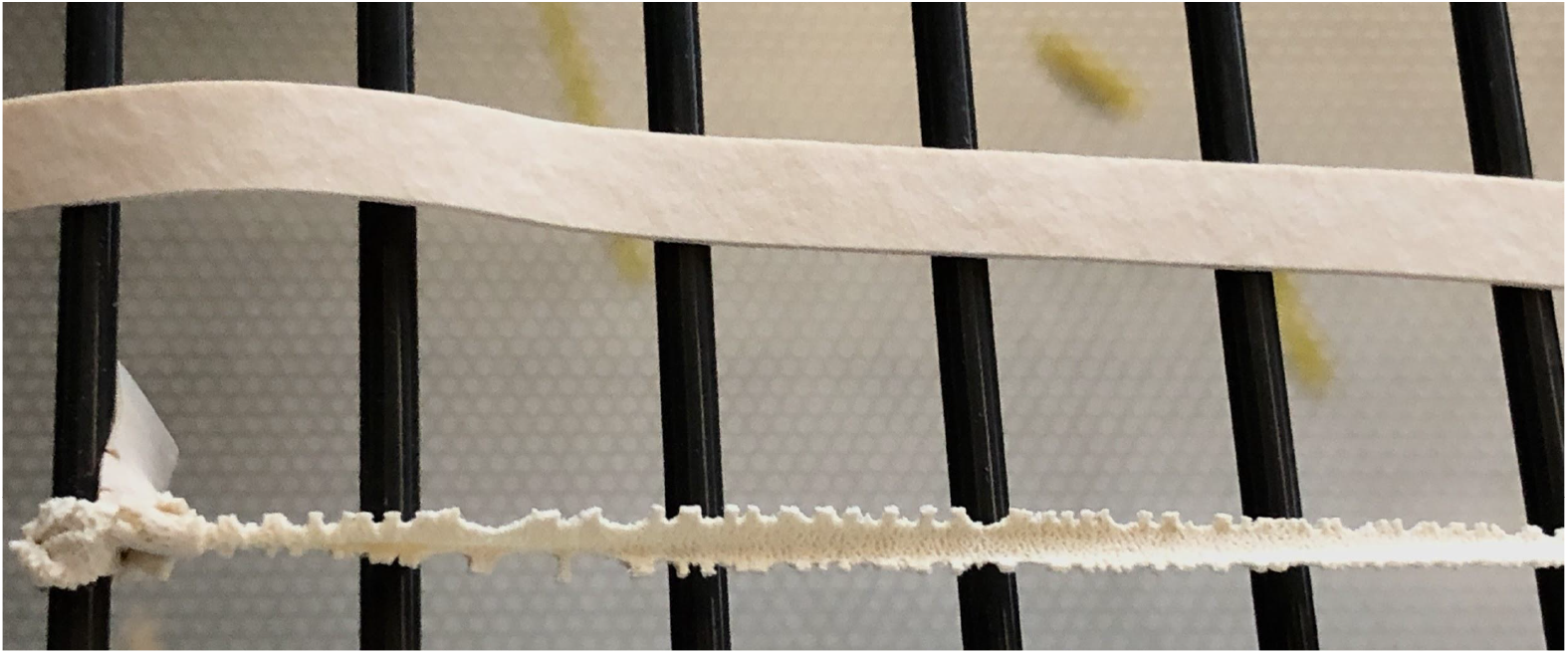
Example of strain-induced ozone damage (20 ppm, 30 minutes, 24°C, 38% RH) to bands separated from a Sperian N1125 respirator. (*Top*) Relaxed (not stretched) band underwent no visible damage and remained functional; (*bottom*) band tied off at 2.4 times its relaxed length during ozone exposure failed.

**Figure S4.**
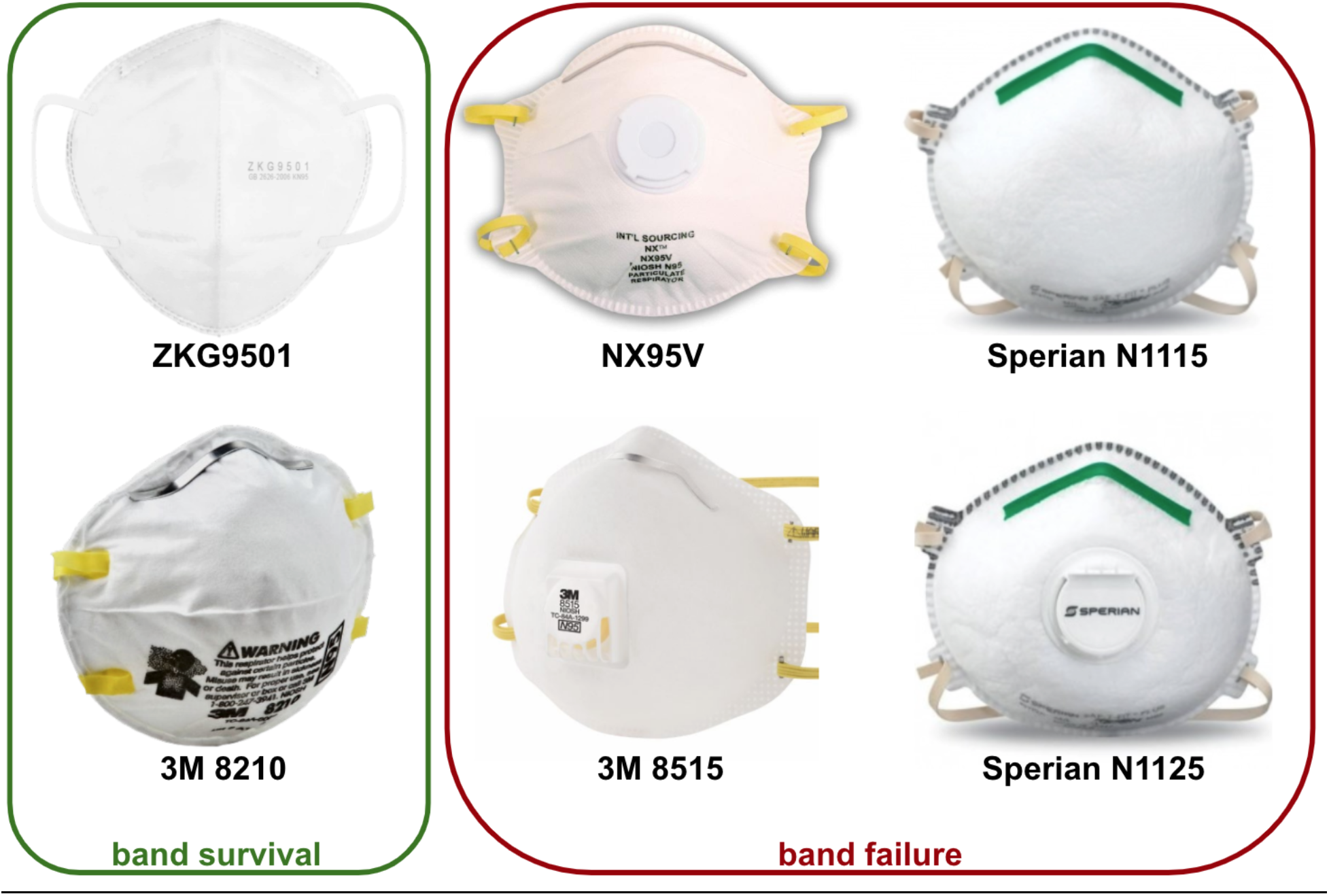
Respirators assessed for headband compatibility with ozone disinfection. All of the respirators with failed bands feature stapled attachments.

**Figure S5.**
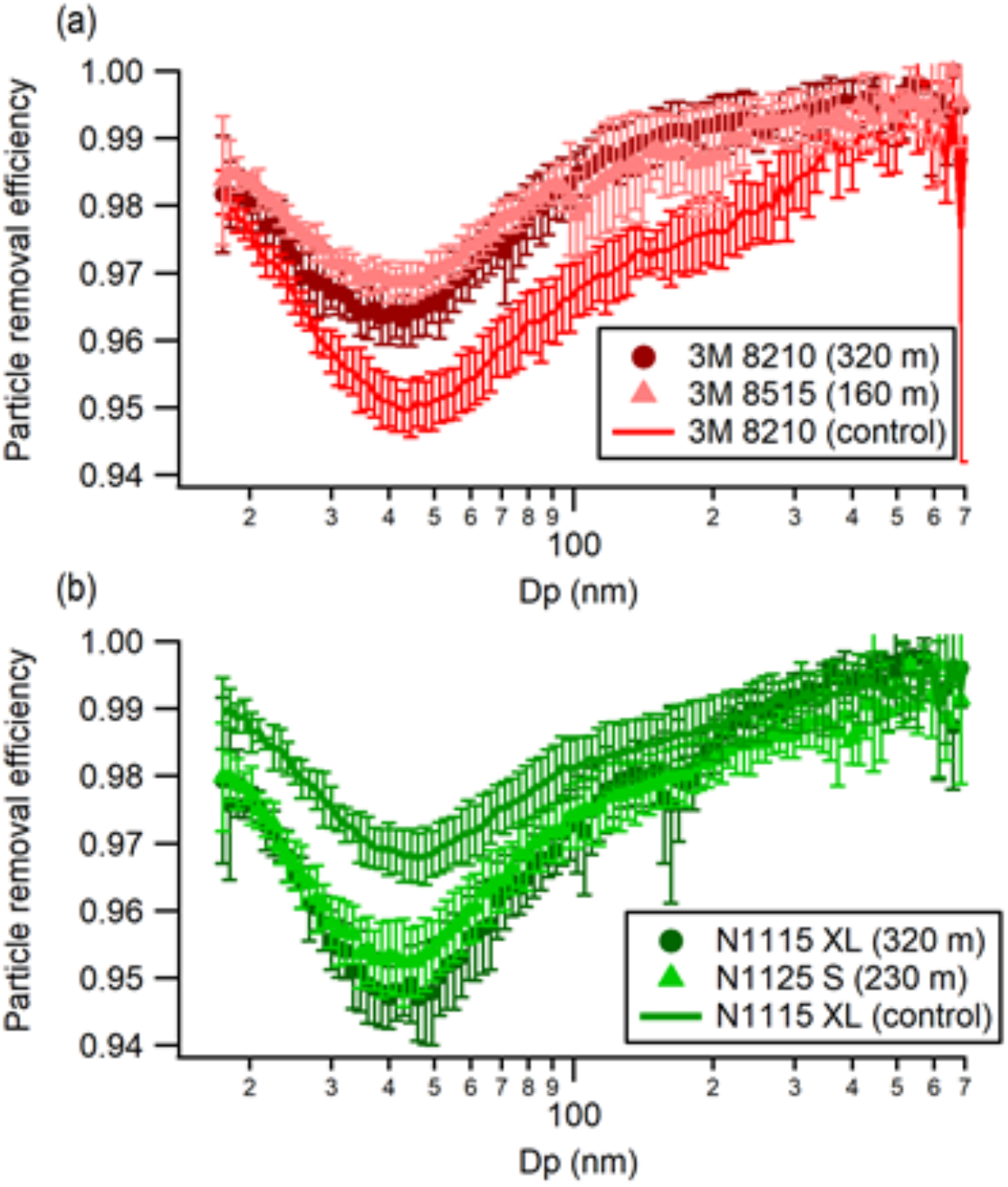
Particle filtration efficiency of untreated (control) and ozone-treated N95 respirators. Uncertainties are calculated from one standard deviation of aerosol volume concentration measured by the SMPS instrument. a) 3M 8210 and 8515 respirators. b) Sperian N1115 XL and N1125 S respirators.

